# Bicycle Helmet Influence in the New Millennium on United States Head, Traumatic Brain Injury, Upper and Lower Body Injury Rates

**DOI:** 10.1101/2023.02.12.23285812

**Authors:** Chris Gillham

## Abstract

This study compares cycling participation and helmet wearing survey results with bicycle-related total injuries, head injuries, traumatic brain injuries and upper/lower body injuries for all-ages, youth and adults who presented to emergency departments or were hospitalized in the United States from 2001 to 2020. Demographic increases/decreases in bicycle-related total injuries concur with participation survey trends, and this is reflected through injury trends for body parts not influenced by helmet wearing. The decrease in 0-17yo total ED presentations from 2001-2010 to 2011-2020 was greater than the decrease in head injuries but the decrease in total hospital admissions was less than the decrease in hospitalized head injuries. The TBI proportion of 0-17yo head injury ED presentations increased significantly, and to a lesser extent among hospitalized head injuries. 18yo+ head injury ED presentations increased at a significantly greater rate than total injuries from 2001-2010 to 2011-2020. 18yo+ hospital admissions more than doubled and head injuries increased 81.6%. The TBI proportion among 18yo+ ED head injuries increased by 9%, while the hospitalized head injury TBI proportion increased from 84.2% to 86.8%. Future studies should examine the relationship between cycling participation and head injuries to determine why total youth injuries including head injury declines were commensurate with participation declines, why adult total injuries including head injuries increased at rates above estimated participation trends, and why the TBI proportion of head injuries has increased despite a greater number of cyclists wearing helmets since 2001.

## 1.1. Introduction

The ongoing global debate about the efficacy of mandatory bicycle helmet legislation largely focuses on claims of reduced head injury risk (1, 2, 3, 4), the legislative influence on cycling participation (5, 6, 7, 8, 9) and/or the likelihood of cyclists crashing because of risk compensation when wearing a helmet (10, 11, 12, 13, 14, 15, 16).

In Australia and New Zealand, the first two countries to enforce all-age bicycle helmet laws in the 1990s, cyclist hospitalized injury numbers have increased significantly despite surveys showing a decline in cycling participation.

Thirty years after enforcement of all-age helmet laws in all Australian states and territories, public opposition remains and in 2018 the country’s largest cycling organisation, the Bicycle Network, reversed its long-standing support for such laws with a recommendation for partial adult repeal. Few countries have mimicked Australia’s all-age laws and over the past decade several countries (Israel, Malta, Bosnia and Herzegovina) have either fully or partially repealed all-age helmet laws, their governments mostly citing evidence that they discourage cycling.

This comprehensive analysis of United States cyclist emergency department (ED) and hospital admission injury data aims to clarify total, upper and lower body, head and traumatic brain injury rates from 2001 to 2020, comparing youth and adult trends, cycling participation and helmet wearing survey data. The database of the National Electronic Injury Surveillance System-All Injury Program (NEISS-AIP) administered by the US Consumer Product Safety Commission (17) is used to calculate the numbers and annual proportionate changes in total injuries, head injuries, traumatic brain injuries (TBI), and upper and lower body injuries among all-ages as well as 0-17yo and 18yo+ cyclists from 2001 to 2020. Decadal comparisons are made between 2001-2010 and 2011-2020 in each category.

Body injuries apart from the head, particularly lower body injuries, are in no way influenced by the wearing of a helmet during a bicycle crash and can be considered a proxy gauge mostly of participation but also overall road safety influenced by traffic laws including speed limits and blood alcohol limits, road user safety education and road or cycling infrastructure. Various studies have found reductions in head injury risk from helmet wearing ranging from 15% to 85% (18, 3). Some studies have found declines in participation following law enforcement that may counteract injury benefits by damaging public health as a result of fewer people riding bikes and instead driving motor vehicles with consequent increases in CO2 emissions and greater crash risk from increased traffic density (5, 6, 7, 8, 9).

### 1.2. Helmet law influence

In America there are 21 states that have mandatory helmet laws for children and teenagers, as well as the District of Columbia and numerous local municipalities. School regulations and parental rules also demand helmet use by youth in non-compulsory state jurisdictions and they are worn voluntarily by many adults across America. A majority of US jurisdictions that have mandated bicycle helmets for children and teenagers did so during the late 1980s and 1990s, with youth helmet law enforcement from 2000 to 2007 in Hawaii, Louisiana, New Hampshire, New Mexico, North Carolina and the District of Columbia.

Published studies by the Centers for Disease Control and Prevention (2011) (19), Chen et al (2013) (20), Coronado et al (2015) (21) and Sarmiento et al (2021) (22) use NEISS-AIP data to calculate the epidemiology and demographic incidence of TBI, all classifying TBI as an NEISS-AIP body part of head only (NEISS disposition code 75) with diagnoses of both concussion (NEISS disposition code 52) and internal organ injury (NEISS disposition code 62). Concussions and internal organ injury to the head are also used in this study to identify TBI. The CDC, Chen et al, Coronado et al and Sarmiento et al studies considered only ED presentations whereas this study also considers hospital admissions, both for comparison and as a proxy indicator of injury severity. Also, these authors base their findings on incidence per 100,000 population instead of estimated cyclist numbers based on participation surveys.

Census population data is more accurate than participation survey data but the latter is considered in this study to be a more reliable representation of injuries per participating cyclist. Population demographic proportions have changed with an ageing population but have consistently increased since 2000, whereas cycling participation surveys show a decrease among youth and an increase among adult. Injuries per 100,000 population are useful but injuries per cyclists participating is more accurate and meaningful, although this study does use a combination of population and participation ratios.

In 2013, Chen et al found among those aged 0 to 14 years, the greatest incidence of ED injuries from 2001 to 2008 occurred to the face, head, hand, arm and wrist. For those aged 15 to 64 years, the upper trunk, face and head were the most injured body parts. For those aged 65 years and older, the head, upper trunk and lower trunk had the highest incidence of injury. Overall, the body parts comprising the largest proportions of injury were the face (15.6%), head (12.6%) and upper trunk (12.1%). The top three injured body parts when the injury involved a motor vehicle were the head (15.2%), upper trunk (14.5%) and face (13.5%). Chen et al found that from 2001 to 2008, children aged 10 to 14 years represented the age group with the largest proportion of ED bicycle injuries, although these injuries were less likely to stem from a motor vehicle collision than the injuries suffered by cyclists aged 15 years and older.

Coronado et al found the NEISS-AIP database showed rates of ED-treated sports and recreation related TBI increased 62% during 2001-2012, mostly affecting the 0-19yo demographic. Males in all-age groups had the highest number of traumatic brain injuries sustained through bicycling. Alfrey et al (2020) (23) found cyclists wearing helmets were less likely to sustain a serious head injury, skull fracture or facial fractures compared to riders without helmets. However, helmets did not prevent concussions, the most common injury among patients in a bicycle crash. In their 2021 study, Sarmiento et al found there were an estimated 596,972 ED visits involving bicycle-related TBIs from 2009 to 2018, with a 27.7% decrease in the incidence per 100,000 population, a 48.7% decrease among children aged 0-17yo and a 5.5% decrease among adults.

### 1.3. Helmet wearing rates

US bicycle helmet wearing surveys are scarce but assessments of known data suggest a significant increase in wearing rates since the 1990s. The US Consumer Product Safety Commission (1995) (24) found that in 1991 about 17.6% of all cyclists, 14.7% of 0-20yo cyclists and 21.5% of 20yo+ cyclists wore helmets all or most of the time.

Dellinger and Kresnow (2010) (25) found that in 2001-2002, 48% of children aged 5-14 years in the US always wore a helmet, 23% sometimes wore a helmet and 29% never wore a helmet. They estimated the proportion of children who always wore a helmet increased from 25% in 1994 to 48% in 2001-2002.

More recent analysis suggests that about 63% of youth and 46% of adults regularly wore a helmet when cycling in 2019 (26, 27, 28), and a 2019 national survey reported 46% of all cyclists never wore a helmet (29). There is general consensus that youth helmet wearing rates are higher than adult helmet wearing rates, and that wearing rates among all ages increased from 2001 to 2020 either through mandate or personal choice.

### 1.4. Cycling participation

US Census Bureau data sourced from the National Sporting Goods Association (NSGA) show a 42.5% decline in 7-17yo cycling participation from 1995 to 2009 (30). NSGA survey data (31) show 17.6 million Americans aged 7-17yo cycled at least once a year in 2000 and 10.1 million in 2014 - a 42.6% reduction. 18y+ cycling was static at 25.5 million in 2000 and in 2014.

National Household Travel Survey (NHTS) data (32) for bicycle trips in 1995, 2001, 2009 and 2017 suggest the demographic cycling shift may be generational, with people continuing to ride bicycles as they age - including the demographic bulge of baby boomers - but the growth in adult cycling countered by a significant decline in children and teenagers riding bikes

**Table 1:** National Household Travel Survey bicycle trip estimates for America (per 100,000 population)

| **Age** | **1995** | **2001** | **2009** | **2017** |
| --- | --- | --- | --- | --- |
| **5-15yo** | 48,012 | 41,029 | 35,297 | 16,398 |
| **16+** | 6,594 | 6,614 | 10,310 | 10,865 |

Gillham and Rissel (2015) (33) collated data from the US Census Bureau, the Outdoor Industry Association (OIA) and the NSGA to find cycling participation at least once per year among 7-17 year old children in the US declined 23.1% from a 1995-2003 average of 18,593,000 to a 2004-2012 average of 14,296,889. Over the same time period, 7-17 year old cyclist all-body injuries in the US declined from a 1995-2003 average of 291,970 to a 2004-2012 average of 222,869, a 23.7% decline, and concussions decreased by 2.1%.

Participation surveys by the OIA (34) suggest annual average cycling by 6-17yo youth on all surfaces decreased by 12.6% (13,697,571 to 11,174,714) and on road/paved surfaces by 18.4% (14,677,429 to 12,826,000) from 2006-2012 to 2013-2019. OIA data suggest cycling participation by the 18yo+ demographic increased on all surfaces by 22.2% (27,362,429 to 33,432,857) and on road/paved surfaces by 9.9% (25,524,000 to 28,047,143) from 2006-2012 to 2013-2019. Participation on all surfaces among 0-17yo was 17,463,000 in 2006 and 12,743,000 in 2019 (−27.0%) while participation on all surfaces among 18yo+ was 22,225,000 in 2006 and 36,140,000 in 2019 (+62.6%).

In 2020, Buehler et al (35) concluded that cycling rates in the US remained unchanged from 2001 to 2017, with a decline in 5-15yo bike riding (10 minutes per day: 4.5% in 2001, 1.9% in 2017 / 20 minutes per day: 3.2% in 2001, 1.3% in 2017 / 30 minutes per day: 2.4% in 2001, 0.9% in 2017) and an increase in the 16-44yo demographic (averaged 10 minutes per day: 1.05% in 2001, 1.5% in 2017 / averaged 20 minutes per day: 0.85% in 2001, 1.2% in 2017 / averaged 30 minutes per day: 0.65% in 2001, 1.0% in 2017).

It is generally accepted that youth cycling in the US has been in decline for the past three decades, countered by an increase in adult cycling. Regular recreational cycling has traditionally been among the most popular exercise routines of youth and there are likely health implications as these are the formative years of muscle development avoiding fat formation as people age, particularly relevant as the US has the highest levels of adult obesity among all developed countries (36). Helmet wearing rates have increased among all-ages from 2001 to 2020, and this study compares the demographic shift in cycling participation with head and other body location injury trends during these 20 years.

## 2.1. Materials and Methods

The web-accessed database of the NEISS-AIP was used, selecting information on patient age, injured body part, principal diagnosis and case disposition for each year from 2001 to 2020 in both the hospitalized and emergency department categories. Only injuries coded as Bicycles & Accessories were used for the analysis. These include bicycles and accessories as well as mountain or all-terrain bicycles and accessories.

ED injuries and hospitalized injuries were collated and calculated as decadal averages for head, TBI, upper and lower body injuries among all-age, 0-17yo and 18yo+ demographics. Head only injuries (NEISS disposition code 75) do not include face (NEISS disposition code 76), eyeball (NEISS disposition code 77), mouth (NEISS disposition code 88), neck (NEISS disposition code 89) or ear (NEISS disposition code 94). TBI was an NEISS-AIP body part of head only (NEISS disposition code 75) with diagnoses of both concussion (NEISS disposition code 52) and internal organ injury (NEISS disposition code 62). Head injuries and TBI were both calculated as a percentage of total ED and/or hospital admissions for each year, and TBI was further calculated as a percentage of brain injuries each year.

Upper and lower body injuries were both calculated as a percentage of total ED and hospital admissions for each year. Lower body injuries were defined as the pubic region (NEISS disposition code 38), lower trunk (NEISS disposition code 79), knee (NEISS disposition code 35), lower leg (NEISS disposition code 36), ankle (NEISS disposition code 37), upper leg (NEISS disposition code 81), foot (NEISS disposition code 83) and toe (NEISS disposition code 93). Upper body injuries were defined as the upper trunk (NEISS disposition code 31), shoulder (NEISS disposition code 30), elbow (NEISS disposition code 32), lower arm (NEISS disposition code 33), wrist (NEISS disposition code 34), upper arm (NEISS disposition code 80), hand (NEISS disposition code 82) and finger (NEISS disposition code 92).

In total, 240 years of different injury data for youth and adults are analysed in this study, among which four years (1.7%) had missing data for hospitalized TBI as the NEISS-AIP does not provide estimates that are below 1,200 or have a Coefficient of Variation exceeding 33% in any given year. Among the 240 years, two years (0.8%) had missing data for hospitalized head injuries, again being less than 1,200 or having a Coefficient of Variation exceeding 33% in those years.

One year among the 240 years of data, or 0.4%, had missing data for hospitalized upper body injuries. One year among the 240, or 0.4%, had missing data for emergency department TBI, again due to either a low number or high CV. If any decade had a missing year or years, their decadal averages rather than totals were compared with the preceding or following decade.

Emergency department patients were categorised as those treated and released, or examined and released without treatment (NEISS disposition code 1). Hospitalised patients were categorised as those treated and transferred (NEISS disposition code 2), treated and admitted for hospitalisation (NEISS disposition code 4), and held for observation (NEISS disposition code 5).

Cycling participation data were sourced from the NSGA, the OIA and the NHTS.

## 3.1. Results

NEISS-API data for ED presentations was collated and decadal totals compared to ascertain trends for less severe injuries to different body parts among all-ages and youth/adult demographics., with averaged results tabulated below.

| All ages annual averages |  |  |  |  |  |  |  |  |  |  |
| --- | --- | --- | --- | --- | --- | --- | --- | --- | --- | --- |
| ED | All injuries | Head injuries | TBI | % head all | % TBI all | % TBI head | Lower body | Upper body | % lower body all | % upper body all |
| 2001-2010 | 401,685 | 59,333 | 37,787 | 14.8% | 9.4% | 63.7% | 143,328 | 199,031 | 35.7% | 49.5% |
| 2011-2020 | 365,986 | 64,607 | 50,913 | 17.7% | 13.9% | 78.8% | 133,646 | 176,074 | 36.5% | 48.1% |
| # change | -35,699 | 5,274 | 13,126 |  |  |  | -9,682 | -22,957 |  |  |
| % change | -8.9% | +8.9% | +34.7% | +20.1% | +48.7% | +24.4% | -6.8% | -11.5% | +2.5% | -2.6% |
| Hospital |  |  |  |  |  |  |  |  |  |  |
| 2001-2010 | 25,502 | 8,646 | 7,063 | 33.9% | 27.7% | 81.7% | 8,595 | 8,269 | 33.7% | 32.4% |
| 2011-2020 | 38,967 | 11,363 | 9,715 | 29.2% | 24.9% | 85.5% | 13,393 | 16,030 | 34.4% | 41.1% |
| # change | 13,465 | 2,717 | 2,651 |  |  |  | 4,798 | 7,761 |  |  |
| % change | +52.8% | +31.4% | +37.5% | -13.3% | -9.1% | +4.9% | +55.8% | +93.9% | +0.3% | +25.4% |
| 0-17yo annual averages |  |  |  |  |  |  |  |  |  |  |
| ED | All injuries | Head injuries | TBI | % head all | % TBI all | % TBI head | Lower body | Upper body | % lower body all | % upper body all |
| 2001-2010 | 233,436 | 37,533 | 22,287 | 16.1% | 9.5% | 59.3% | 84,015 | 111,889 | 36.0% | 47.8% |
| 2011-2020 | 156,545 | 27,779 | 20,667 | 17.8% | 13.2% | 74.6% | 59,146 | 71,823 | 37.9% | 46.1% |
| # change | -76,891 | -9,753 | -1,620 |  |  |  | -24,868 | -40,066 |  |  |
| % change | -32.9% | -26.0% | -7.3% | +10.6% | +38.5% | +25.8% | -29.6% | -35.8% | +5.3% | -3.6% |
| Hospital |  |  |  |  |  |  |  |  |  |  |
| 2001-2010 | 10,903 | 3,872 | 3,044 | 35.5% | 27.9% | 78.6% | 3,153 | 3,878 | 28.8% | 35.7% |
| 2011-2020 | 8,099 | 2,460 | 2,076 | 30.4% | 25.6% | 84.4% | 2,572 | 3,313 | 31.7% | 41.2% |
| # change | -2,804 | -1,412 | -968 |  |  |  | -581 | -565 |  |  |
| % change | -25.7% | -36.5% | -31.8% | -14.9% | -9.3% | +3.6% | -18.4% | -14.6% | +10.1% | +15.4% |
| 18yo+ annual averages |  |  |  |  |  |  |  |  |  |  |
| ED | All injuries | Head injuries | TBI | % head all | % TBI all | % TBI head | Lower body | Upper body | % lower body all | % upper body all |
| 2001-2010 | 168,249 | 21,801 | 15,946 | 13.0% | 9.5% | 73.1% | 59,313 | 87,136 | 35.2% | 52.0% |
| 2011-2020 | 209,441 | 36,828 | 30,241 | 17.6% | 14.4% | 82.1% | 74,499 | 104,249 | 35.6% | 49.9% |
| # change | 41,192 | 15,027 | 14,295 |  |  |  | 15,186 | 17,114 |  |  |
| % change | +24.5% | +68.9% | +89.6% | +37.6% | +56.2% | +15.6% | +25.6% | +19.6% | +1.1% | -4.0% |
| Hospital |  |  |  |  |  |  |  |  |  |  |
| 2001-2010 | 14,599 | 4,889 | 4,114 | 33.5% | 28.2% | 84.1% | 5,443 | 4,490 | 37.9% | 29.7% |
| 2011-2020 | 30,868 | 8,877 | 7,709 | 28.8% | 25.0% | 86.8% | 10,821 | 12,709 | 34.5% | 40.2% |
| # change | 16,269 | 3,988 | 3,595 |  |  |  | 5,378 | 8,219 |  |  |
| % change | +111.4% | +81.6% | +87.4% | -10.2% | -3.8% | +4.2% | +98.8% | +183.0% | -9.0% | +35.4% |

### 3.2. Emergency Department head injuries

NEISS-AIP estimates for all-age cyclist injuries show an overall reduction of 356,988 total injuries presenting at US hospital emergency departments from 2001-2010 to 2011-2020, but a 52,739 increase in head injuries and a 131,256 increase in TBI. As a proportion of total injuries presenting to ED between these decades, head injuries increased 20.1% and TBI increased 48.7%. As a proportion of head injuries presenting to ED, TBI increased 24.4% from 2001-2010 to 2011-2020 (Appendix Table 2).

Among the 0-17yo demographic there was an overall reduction of 768,906 total injuries presenting at US hospital emergency departments from 2001-2010 to 2011-2020, including a 97,533 reduction in head injuries and a 16,203 reduction in TBI. As a proportion of total injuries presenting to ED between these decades, 0-17yo head injuries increased 10.6% and TBI increased 38.5%. As a proportion of head injuries presenting to ED, TBI increased 25.8% (Appendix Table 3).

Among the 18yo+ demographic there was an overall increase of 411,921 total injuries presenting at US hospital emergency departments from 2001-2010 to 2011-2020, including a 150,271 increase in head injuries and a 158,896 increase in TBI. As a proportion of total injuries presenting to ED between these decades, 18yo+ head injuries increased 37.6% and TBI increased 56.2%. As a proportion of head injuries presenting to ED, TBI increased 15.6% (Appendix Table 4).

### 3.3. Emergency Department non-head injuries

Among all ages there was a 356,988 decrease in cyclist total injuries, including a 96,820 decrease in lower body injuries and a 229,571 decrease in upper body injuries, presenting to US hospital emergency departments between 2001-2010 and 2011-2020. As a proportion of total injuries presenting to ED between these decades, all-age lower body injuries increased 2.5% and upper body injury presentations decreased 2.6% (Appendix Table 5).

Among the 0-17yo demographic there was a 768,906 decrease in cyclist total injuries, including a 248,683 decrease in lower body injuries and a 400,661 decrease in upper body injuries, presenting to US hospital emergency departments between 2001-2010 and 2011-2020. As a proportion of total injuries presenting to ED between these decades, 0-17yo lower body injury presentations increased 5.3% and upper body injury presentations decreased 3.6% (Appendix Table 6).

Among the 18yo+ demographic there was a 411,921 increase in cyclist total injuries, including a 151,862 increase in lower body injuries and a 171,139 increase in upper body injuries, presenting to US hospital emergency departments between 2001-2010 and 2011-2020. As a proportion of total injuries presenting to ED between these decades, 18yo+ lower body injury presentations increased 1.1% and the proportion of upper body injuries decreased 4.0% (Appendix Table 7).

### 3.4. Hospital admission head injuries

NEISS-API data for hospital admissions was collated and decadal totals compared to ascertain trends for more severe injuries to different body parts among all-ages and youth/adult demographics.

NEISS-AIP estimates for all-age cyclist injuries show a 134,651 increase in total injuries admitted to US hospitals from 2001-2010 to 2011-2020, including a 27,174 increase in head injuries and a 26,511 increase in TBI. As a proportion of total hospitalized injuries between these decades, head injuries decreased 13.3% and TBI decreased 9.1%. As a proportion of head injuries admitted to hospital, TBI increased 4.9% (Appendix Table 8).

Among the 0-17yo demographic there was a 28,044 decrease in cyclist total injuries admitted to US hospitals from 2001-2010 to 2011-2020, including a 14,121 decrease in head injuries and an 11,757 decrease in TBI. As a proportion of total injuries admitted to hospital between these decades, 0-17yo head injuries decreased 14.9% and TBI decreased 9.3%. As a proportion of head injuries admitted to hospital, TBI increased 3.6% (Appendix Table 9).

Among the 18yo+ demographic there was a 162,693 increase in cyclist total injuries admitted to US hospitals from 2001-2010 to 2011-2020, including a 49,658 increase in head injuries and a 48,295 increase in TBI. As a proportion of total injuries admitted to hospital between these decades, 18yo+ head injuries decreased 10.2% and TBI decreased 3.8%. As a proportion of head injuries admitted to hospital, TBI increased 4.2% (Appendix Table 10).

### 3.5. Hospital admission non-head injuries

Among all ages there was a 134,651 increase in cyclist total injuries admitted to US hospitals from 2001-2010 to 2011-2020, including a 47,976 increase in lower body injuries and a 77,609 increase in upper body injuries. As a proportion of total injuries admitted to hospital between these decades, all-age lower body injuries increased 0.3% and upper body injuries increased 25.4% (Appendix Table 11).

Among the 0-17yo demographic there was a 28,044 decrease in cyclist total injuries admitted to US hospitals from 2001-2010 to 2011-2020, including a 5,811 decrease in lower body injuries and a 5,654 decrease in upper body injuries. As a proportion of total injuries admitted to hospital between these decades, 0-17yo lower body injuries increased 10.1% and upper body injuries increased 15.4% (Appendix Table 12).

Among the 18yo+ demographic there was a 162,693 increase in cyclist total injuries admitted to US hospitals from 2001-2010 to 2011-2020, including a 53,780 increase in lower body injuries and an 86,675 increase in upper body injuries. As a proportion of total injuries admitted to hospital between these decades, 18yo+ lower body injuries decreased 9.0% and upper body injuries increased 35.4%. (Appendix Table 13)

## 4.1. Discussion

The NEISS-AIP data suggest there was an annual average 8.9% decrease in all-age US hospital emergency department cyclist presentations for total injuries from 2001-2010 to 2011-2020, but a 52.8% increase in hospital admissions. With a decline in the number of cyclists presenting to ED for mostly minor injuries but an increase in the number either transported by ambulance or transferred from ED for hospital admission, this indicates an increase in injury severity.

There was an 8.9% annual average increase in all-age cyclist hospital emergency department presentations for head injuries from 2001-2010 to 2011-2020, and a 31.4% increase in hospital admissions for head injuries. All age cyclist hospital emergency department presentations for TBI increased 34.7% from 2001-2010 to 2011-2020, and hospital admissions for TBI increased 37.5%.

The 8.9% head and 34.7% TBI increases in annual average ED presentations, compared to the 8.9% decrease in total ED injuries, represent a proportionate increase of 20.1% in head injuries and 48.7% increase in TBI for patients not admitted to hospital. However, the 31.4% head and 37.5% TBI increases in hospital admissions are less than the 52.8% increase in total injury admissions, representing a proportionate decrease of 13.3% in head injuries and a 9.1% decrease in TBI.

Among cyclists with head injuries, the annual average proportion of emergency department presentations with TBI increased by 24.4% from 2001-2010 to 2011-2020, and the proportion of hospital admissions with TBI increased by 4.9%. TBI represented 63.7% of the average annual number of all-age ED head injuries in 2001-2010 (59,333/37,787) and 78.8% of all-age ED head injuries in 2011-2020 (64,607/50,913). TBI represented 81.7% of the average annual number of all-age hospitalized head injuries in 2001-2010 (8,646/7,063) and 85.5% of all-age hospitalized head injuries in 2011-2020 (11,363/9,715).

### 4.2. Helmet efficacy

These results raise questions about the efficacy and possible drawbacks of bicycle helmets which over the two decades saw an increase in the number of US cyclists of all ages wearing them, mostly through laws among youth and voluntary wearing among adults.

It is possible that TBI increased proportionally because of a decrease in external head wounds such as fractures. However, the NEISS has only two years that estimate cyclist head fractures presenting to ED (5,264 in 2019, 6,999 in 2020) and three years estimating cyclist head fractures admitted to hospital (1,392 in 2012, 5,764 in 2019, 6,445 in 2020). In all other years, it’s likely there were either less than 1,200 cases or the Coefficient of Variation exceeded 33% so that no estimates are returned.

The 8.9% increase in all-age ED head injuries and 31.4% increase in all-age hospitalized head injuries, compared to the 34.7% ED increase in TBI and 37.5% hospitalized increase in TBI from 2001-2010 to 2011-2020, along with fractures seemingly increasing in the 2011-2020 decade, suggests TBI did increase as a greater proportion of total head injuries suffered by cyclists. The available data show ED skull fractures increased 33.0% and hospitalized skull fractures increased 11.8% from 2019 to 2020, whereas ED presentations for TBI increased 12.9% and hospitalized TBI increased 8.3%. These years overlap the COVID-19 pandemic and associated lockdowns resulting in increased cycling and reduced commuter motor vehicle traffic. The increased TBI proportion of all-age head injuries from 2001-2010 to 2011-2020 coincided with increased helmet wearing among cyclists and this does not indicate they have reduced the risk of intracranial brain injury, possibly due to their wider head impact area accelerating rotational brain injury (37).

Among all ages there were an annual average 35,699 fewer US emergency department total injury presentations, 5,274 more head injury presentations and 13,126 more TBI presentations in 2011-2020 than in 2001-2010. Among all ages there were an annual average 13,465 more total injury hospital admissions, 2,717 more head injury admissions and 2,651 more TBI admissions in 2011-2020 than in 2001-2010.

From 2001-2010 to 2011-2020 there was an annual average decrease of 9,682 all-age lower body injuries and 22,957 fewer upper body injuries presenting to emergency departments. However, there was an annual average increase of 4,798 all-age lower body injuries and 7,761 upper body injuries admitted to hospital.

The larger decrease in upper than lower body injury ED presentations differs with claims that bicycle helmets increase the odds of upper body injury, but the smaller increase in lower than upper body hospital admissions suggests an increase in upper body injury severity.

An analysis of 0-17yo and 18yo+ injuries allows a comparison of youth demographics with higher helmet wearing incidence but reduced cycling participation, and adult demographics with lower helmet wearing incidence but increased cycling participation.

### 4.3. 0-17yo injuries

The 0-17yo cyclist demographic had an annual average 76,891 fewer total injury ED presentations from 2001-2010 to 2011-2020, a 32..9% decrease which may be indicative of helmet protection, reduced youth cycling participation and/or improved vehicle traffic regulations and infrastructure that are beneficial to the safety of children and teenagers riding bicycles.

There were 9,753 fewer head injuries but this 26.0% decrease was less than the 32.9% decrease in total injuries, suggesting that helmet protection was not a significant influence. However, 0-17yo total injury hospital admissions declined by an annual average 2,804, or 25.7%, while head injury hospital admissions declined by 1,412, or 36.5%, suggesting helmets reduced head injury severity. Head injuries represented 35.5% of total injury hospital admissions in 2001-2010 (10,903/3,872) and 30.4% in 2011-2020 (8,099/2,460).

TBI represented 59.3% of 0-17yo ED head injuries in 2001-2010 (37,533/22,287) and 74.6% of 0-17yo ED head injuries in 2011-2020 (27,779/20,667). TBI represented 78.6% of 0-17yo hospitalized head injuries in 2001-2010 (3,872/3,044) and 76.0% of 0-17yo hospitalized head injuries in 2011-2020 (2,460/2,076).

0-17yo annual average lower body injury ED presentations decreased by 24,868, or 29.6%, from 2001-2010 to 2011-2020, while upper body injuries decreased by 40,066, or 35.8%. Lower body injuries are not influenced by helmet wearing except for risk compensation possibly increasing crash frequency, and the non-head injury data suggest a participation decline commensurate with cycling survey numbers.

The greater number and percentage decrease in upper body injuries, while suggesting reduced participation, does not support claims that helmets increase upper body injury risk - particularly among youth who have a higher helmet wearing proportion than adults. However, this contention is not supported by NEISS-AIP

0-17yo hospital admission data showing lower body injuries decreased by an annual average 581 from 2001-2010 to 2011-2020, an 18.4% decrease, while upper body injuries decreased 565, or 14.6%. The data for hospitalized cyclists also suggest that upper body injuries have played a greater role than lower body injuries in replacing the lower proportionate share of head injuries:

**2001-2010** head : 35.5% (10,903/3,872); lower body : 28.9% (10,903/3,153); upper body : 35.6% (10,903/3,878)

**2011-2020** head : 30.4% (8,099/2,460); lower body : 31.8% (8,099/2,572); upper body : 40.9% (8,099/3,313)

The decline in 0-17yo total hospitalized injuries should be compared with estimates of youth cycling participation, with OIA surveys suggesting average annual cycling by 6-17yo youth decreased 12.6% on all surfaces and by 18.4% on road/paved surfaces from 2006-2012 to 2013-2019. Participation among 0-17yo on all surfaces declined 27.0% from the year 2006 to the year 2019.

NSGA surveys suggest a 42.6% reduction in 7-17yo cycling at least once a year from 2000 to 2014 (17.6 million to 10.1 million), and NHTS surveys suggest 5-15yo cycling per 100,000 population decreased 60.0% from 2001 to 2017 (41,029 to 16,398) (38).

This compares with a 32.9% decrease in total 0-17yo ED presentations and a 26.0% decrease in head injuries from 2001-2010 to 2011-2020, with a 25.7% decrease in total hospital admissions and a 36.5% decrease in hospitalized head injuries. It is likely the reduction in youth total injuries and head injuries was due to an equal or similar reduction in youth cycling participation.

Comparing OIA participation and NEISS injury figures for the 6-17yo demographic from 2006-2012 to 2013-2019, total injuries per 100,000 bicycle riders declined 16.8% and head injuries per 100,000 riders declined 22.7% (Appendix Table 15).

### 4.4. 18yo+ injuries

The 18yo+ cyclist demographic had an annual average 41,192 more total injury ED presentations in 2011-2020 than 2001-2010, a 24.5% increase which may be indicative of increased adult cycling participation.

OIA surveys suggest that within the 18yo+ adult demographic, cycling on all surfaces increased 22.2% and on road/paved surfaces by 9.9% from 2006-2012 to 2013-2019, with a 62.6% increase from the year 2006 to the year 2019. NSGA survey data suggest 18yo+ cycling participation was unchanged between those years at 25.5 million. NHTS data show that from 2001 to 2017 among the 16yo+ demographic there was a 64.3% increase (6,614 to 10,865) in cycling per 100,000 population.

There were an annual average 15,027 more 18yo+ head injuries from 2001-2010 to 2011-2020, and this 68.9% increase was substantially more than the 24.5% increase in total injuries, despite a greater proportion of adult cyclists wearing helmets in 2011-2020 than 2001-2010. However, 18yo+ total injury hospital admissions increased by an annual average 16,269, or 111.4%, while head injury hospital admissions increased by 3,988, or 81.6%, suggesting helmets reduced head injury severity. Head injuries represented 33.5% of 18yo+ annual average total injury hospital admissions in 2001-2010 (14,599/4,889) and 28.8% in 2011-2020 (30,868/8,877).

TBI represented an annual average 73.1% of 18yo+ ED head injuries in 2001-2010 (15,946/21,801) and 82.1% in 2011-2020 (36,828/30,241). TBI represented 84.1% of 18yo+ hospitalized head injuries in 2001-2010 (4,889/4,114) and 86.8% in 2011-2020 (8,887/7,709).

Annual average 18yo+ lower body injury ED presentations increased by 15,186, or 25.6%, from 2001-2010 to 2011-2020, while upper body injuries increased by 17,114, or 19.6%. With lower body injuries not influenced by helmet wearing, their 25.6% increase probably reflects increased adult cycling participation. Again, the greater percentage increase in lower than upper body injuries does not support claims that helmets increase upper body injury risk.

However, NEISS-AIP 18yo+ hospital admission estimates show annual average lower body cyclist injuries increased 5,378 from 2001-2010 to 2011-2020, a 98.8% increase, while upper body injuries increased 8,219, or 183.0%. With the proportion of adult helmet wearing increasing from 2001-2010 to 2011-2020, this supports claims that helmets increase the risk of upper body injuries and also suggests increased upper body injury severity.

The estimates for hospitalized cyclists also suggest that the upper body injury proportion of total 18yo+ total injuries has increased significantly while the lower body proportion has declined:

**2001-2010** head : 33.5% (14,599/4,889); lower body : 37.3% (14,599/5,443); upper body : 30.8% (14,599/4,490)

**2011-2020** head : 28.8% (30,868/8,877); lower body : 35.1% (30,868/10,821); upper body : 41.2% (30,868/12,709)

Comparing OIA participation and NEISS injury figures for the 18yo+ demographic from 2006-2012 to 2013-2019, total injuries per 100,000 bicycle riders increased 33.3% and head injuries per 100,000 riders increased 16.2%. (Appendix Table 16)

Alternatively comparing NHTS participation and NEISS injury figures for different demographics in 2001 and 2017: total injuries among 5-15yo per 100,000 bicycle riders increased 47.6% and head injuries per 100,000 riders increased 8.6%; total injuries among 16yo+ per 100,000 bicycle riders increased 62.9% and head injuries per 100,000 riders increased 67.9% (Appendix Table 17).

These results are similar to a 2019 study in which Kirkpatrick et al (41) compared NEISS data with NSGA participation survey results within the 18yo+ demographic, finding per participant injuries rose from 701/100,000 in 1997 to 1,164/100,000 in 2013, a 66.0% increase, concluding that there was an increased risk of injury for younger riders but injuries among older riders increased due to greater participation.

### 4.5. Analysis

The percentage decline in 0-17yo head injuries in emergency departments is 6.9% less than the decline in total injuries from 2001-2010 to 2011-2020, and the percentage decline in TBI is 25.6% less. The decline in 0-17yo head injury hospital admissions is 10.8% more than the decline in total injuries, and the decline in TBI is 6.1% more.

However, the 18.4% decline in 0-17yo hospitalized lower body injuries and 14.6% decline in hospitalized upper body injuries suggest a majority of all body part injury trends are due to reduced youth cycling participation. The greater proportion of hospitalized upper body injuries in 2011-2020 than 2001-2010 also suggests a change in youth bicycle crash characteristics. The 25.8% increase in the 0-17yo TBI proportion of head injuries in ED from 2001-2010 to 2011-2020 suggests greater intracranial injury risk, partly supported by the 3.6% increase in 0-17yo TBI proportion of hospitalized head injuries.

The increase in 18yo+ head injuries in emergency departments is 44.4% more than the increase in total injuries from 2001-2010 to 2011-2020, and the increase in TBI is 65.1% more. The increase in 18yo+ head injury hospital admissions is 29.8% less than the increase in total injuries, and the increase in TBI is 24.0% less. A 25.6% increase in 18yo+ lower body injuries and 19.6% increase in upper body injuries presenting to ED suggests a majority of all body part injury trends are due to increased adult cycling participation. The significantly greater proportion of hospitalized 18yo+ upper body injuries in 2011-2020 than 2001-2010 also suggests a change in adult bicycle crash characteristics.

The 15.6% increase in the 18yo+ TBI proportion of head injuries in ED from 2001-2010 to 2011-2020 suggests greater intracranial injury risk, as does the 4.2% increase in the 18yo+ TBI proportion of hospitalized head injuries.

NEISS-AIP injury data can be interpreted in various ways to estimate increased helmet use influence on head, TBI, upper and lower body injury trends, but the bulk of evidence including participation surveys and non-head injuries suggests an increase in the crash/injury ratio per cyclist but a decrease in head injury severity, and an increase in the risk of TBI among cyclists who suffer a head injury during a crash. This indicates either a change in the US cycling environment or greater risk-taking by cyclists, possibly due to increased helmet wearing, resulting in a smaller proportion of head injuries but a greater total number of head, upper and lower body injuries. The NEISS-AIP data also suggest helmets exacerbate TBI and upper body injuries more among adults than child or teenage cyclists, potentially due to the greater height and body mass of adults.

Confounding factors that may influence cyclist injuries include the Affordable Care Act enforced in 2014 which mandates personal health insurance and restricts the ability of insurers to alter premiums based on the health of the individual, although it’s not know how this might affect the risk compensation of cyclists, with or without helmets.

Total registered vehicles in the US increased 22.4% from 2000 to 2019 (225,821,241 to 276,491,174) (39), potentially increasing the risk of collision with cyclists. However and despite the automobile increase, the number of motor vehicle occupant deaths declined 15.3% from 2000-2009 to 2010-2019 (411,649 to 348,591), the number of motor vehicle occupant injuries declined 7.5% (27,250,000 to 25,218,000), and the number of motor vehicle crashes increased 0.3% (61,014,000 to 61,184,000).

Sports utility vehicles (SUVs) represented 8.6% of US automobile sales in 2020 and the proportion of these vehicles on US roads has increased significantly since 2000. Studies have shown they pose a safety risk to pedestrians because of their higher front ends (40), and it’s likely there is a similar risk to cyclists. In recent years, an increasing number of motor vehicles including SUVs have provided pedestrian detection technology with automatic braking. The decrease in American motor vehicle occupant deaths and injuries over the past 20 years suggests that improved vehicle safety features and tougher drink driving, speed and other laws in different states have improved road safety for motorists, and these improvements should also benefit other road users including cyclists. For example, data from the National Highway Traffic Safety Administration (NHTSA) (42) show the US suffered 13,324 alcohol-impaired traffic fatalities in 2000 and 10,142 in 2019, a 23.9% reduction.

Data from the Web-based Injury Statistics Query and Reporting System (WISQARS) administered by the Centers for Disease Control and Prevention (Appendix Table 14) show the crude rate per 100,000 population of 5-15yo hospitalized cyclist injuries declined 42.4% from 2001-2009 to 2011-2019, with 5-15yo vehicle occupant injuries declining 54.8% and 5-15yo pedestrian injuries declining 25.5%. In the 16yo+ demographic, the crude rate per 100,000 population for hospitalized cyclists increased 80.2%, declined 7.4% for vehicle occupants and increased 55.2% for pedestrians. These trends suggest improved safety for vehicle occupants but reduced safety for other road users. Cyclists aged 5-15yo had a greater injury decline than pedestrians in the same demographic, although all surveys suggest this is related to a significant reduction in youth cycling participation. Among the 16yo+ demographic, cyclists suffered a significantly greater injury increase than pedestrians and at a significantly greater rate than the adult cycling participation increase revealed by surveys. These trends suggest increased helmet wearing by adult cyclists from 2001 to 2019 has had little impact on total injuries when compared to other road users, and raise questions about a possibly increased accident rate per cyclist.

## 5.1. Conclusions

Future studies should analyse cyclist total injuries and head injuries and their association with helmet use in the context of participation among different cyclist demographics, while US hospital emergency department presentation and admission data suggest further examination is required to establish whether helmets increase the risk of traumatic brain injury.

Proxy cycling participation trends can be roughly calculated from NEISS-AIP estimates of injuries to non-head body parts, particularly the lower body where injuries are not influenced by helmet wearing. The decline in total youth injuries is similar to the decline in youth participation, and the accuracy of participation survey estimates is reinforced by the 29.6% reduction in annual average 0-17yo ED lower body injuries from 2001-2010 to 2011-2020. For example, Outdoor Industry Association surveys show 6-17yo cycling participation declined 27.0% from 2006 to 2019.

However, NHTS data suggest a 60.0% decline in 5-15yo cycling per 100,000 population from 2001 to 2017. A decline of this magnitude would suggest a greater decline in youth participation than the decline in ED or hospitalized injuries of all types. If accurate, this would indicate an increased rate of accidents and/or injuries per cyclist.

The reason for the youth cycling participation decline is contentious, with competing beliefs whether it’s caused by helmet laws and parent/school helmet wearing coercion or by competing indoor interests centred on computers and the internet. However, the NEISS-AIP hospital admission estimates suggest majority helmet use by children and teenage cyclists reduces head injury severity, with annual average total 0-17yo hospitalized injuries declining by 25.7% from 2001-2010 to 2011-2020 but head injuries declining by 36.5% and lower body injury hospital admissions only declining by 18.4%.

The annual average head injury proportion of total 0-17yo hospital admissions decreased 14.9% from 2001-2010 to 2011-2020 but the upper body proportion decreased less than the lower body proportion, suggesting a slight change in body area injury risk when children and teenagers crash their bicycles.

Within emergency departments, these results were effectively reversed for 18yo+ cyclist injuries due to increased participation, with annual average total ED injury presentations up 24.5% from 2001-2010 to 2011-2020, similar to the lower body injury increase of 25.6%, but head injuries increasing 68.9%.

However, 18yo+ annual average hospital admissions increased 111.4%, considerably more than survey estimates of increased adult cycling participation.. For example, OIA survey data show 18yo+ cycling participation on all surfaces increased 22.2% from 2006-2012 to 2013-2019, or a 62.6% increase from the year 2006 to the year 2019. NHTS data show a 64.3% increase in cycling per 100,000 population among the 16yo+ demographic from 2001 to 2017. This suggests increased injury occurrence and severity among adult cyclists. 18yo+ head injury annual average hospital admissions increased 81.6% from 2001-2010 to 2011-2020, while lower body injuries increased 98.8%. Both are less than the total injury increase but still well above participation estimates.

The annual average 18yo+ head injury proportion of total hospital admissions decreased 4.7% from 2001-2010 to 2011-2020 (33.5% to 28.8%) while the upper body proportion increased 10.4% (30.8% to 41.2%) and the lower body proportion decreased 2.2% (37.3% to 35.1%). This reinforces the suggestion of an increase in upper body injuries possibly related to helmet use. The trend may be influenced by unknown factors such as bicycle design, cycling infrastructure or adult cycling trends of more or fewer people riding in vehicular traffic environments, but adds support to claims that helmet use increases the likelihood of upper body injuries.

A finding of concern is that the TBI proportion of head injured patients increased among all-ages by 24.4% with ED presentations and by 4.9% with hospital admissions from 2001-2010 to 2011-2020. Among 0-17yo, the TBI proportion of head injuries increased 25.8% for ED presentations and 3.6% for hospital admissions. Among 18yo+, the TBI proportion of head injuries increased 15.6% for ED presentations and 4.2% for hospital admissions.

Other studies (3, 4, 22, 23, 36) have found that helmets either reduce or have little influence on TBI. However, this study suggests they increase the likelihood as TBI is estimated as a proportion of head injuries only and isn’t influenced by the number or trends in total injuries, head injuries, population or cycling participation. The finding adds weight to claims that bicycle helmets increase intracranial injury risk due to additional centrifugal force from a wider helmet impact area that increases rotational brain movement when the head suffers a glancing blow during a bicycle crash.

It is also noteworthy that from 2017-2018 to 2019-2020, ED presentations decreased 2.6% for total all-age injuries but increased 14.2% for head injuries and 16.7% for TBI, while hospital admissions increased 16.5% for total all-age injuries, 26.1% for head injuries and 38.0% for TBI, with a possible COVID-19 participation influence in 2020.

Overall, NEISS-AIP injury estimates show little improvement and for adults a probable worsening in head and traumatic brain injuries suffered by US cyclists over a 20 year timespan during which helmet use increased, and further studies should consider relevant cycling participation, helmet wearing and body part injury data when evaluating the efficacy of helmet use whilst cycling. These results suggest claims that child cyclist head injury reductions are indicative of helmet efficacy are not necessarily accurate or mindful of other factors, and should be considered by legislators wishing to mandate bicycle helmets.

The findings of this study raise questions for subsequent studies into the generational shift in US cycling participation, whether this is related to mandatory bicycle helmets for youth, whether there is a consequent impact on public health, whether increased helmet wearing among all generations is causing increased accidents and injuries per cyclist, and whether helmets provide adequate head injury prevention to warrant their mandatory use.

## 6.1 Limitations

Cycling participation estimates are sourced to the US Census Bureau, the National Household Travel Survey, the National Sporting Goods Association and the Outdoor Industry Association. Although each source was roughly in agreement on numbers and trends for all demographics since the 1990s, their accuracy cannot be validated. NHTS estimates of reduced 5-15yo cycling per 100,000 population are particularly large and thus questionable. Helmet wearing estimates are based on the few national surveys conducted since the 1990s. Although calculated averages are unreliable, all sources show an increase in helmet wearing over that time.

NEISS-AIP is based on a nationally representative probability sample of hospitals in the US and its territories, and weighted by the inverse probability of selection to provide national estimates. The NEISS-AIP database includes data from 66 of the 100 NEISS-AIP hospitals, which are representative of the 5,000 hospitals that have a minimum of six beds and a 24-hour ED in the United States and its territories. The NEISS-AIP collects data on nonfatal injuries treated in US emergency departments and each NEISS-AIP case is assigned an inverse probability weight, which represents the inverse of the probability of the case being selected into the sample.

The NEISS-AIP database does not provide patient data on severity of injury or follow-up care. Hospital admissions rather than ED presentations are in this study considered indicative of more severe injuries to the head, lower and upper body. The fact that only bicycling injuries treated in EDs are included within NEISS-AIP implies that the total number of bicycling injuries to different body parts is likely to be an underestimate as injuries treated at home, in urgent care clinics and in GP offices are not reported. The NEISS-AIP is also considered to underestimate the number of TBI cases in hospital emergency departments. The NEISS-AIP does not provide detailed or consistent information on patient use of safety equipment such as helmets, so the effect of helmet use on head injury status could not be assessed.

Due to inadequate data within NEISS-AIP, reliable information on crash/injury locale or motor vehicle involvement is not considered in this study and it is not known if this unreported information could cause bias. NEISS-AIP data consists of the principal diagnosis and primary body part. Some cases for which TBI was a secondary diagnosis, such as skull fractures with an underlying TBI, may not be included. This study does not examine race/ethnicity or socioeconomic differences that may be associated with limited neighbourhood bicycle safety infrastructure that increases or decreases the risk of bicycle-related injuries.

Head injuries in this study exclude face, eyeballs, mouth, neck and ears, with the exception of Table 17. These body parts may be influenced by the presence or otherwise of a helmet in a crash and potentially underestimate any trends in overall head injuries.

This research did not receive any specific grant from funding agencies in the public, commercial or not-for-profit sectors.

## Data Availability

All data produced in the present work are contained in the manuscript

### Appendix

**Table 2:** United States cyclist total injury, head only and traumatic brain injury emergency department presentations, all-ages, 2001-2020. Source: NEISS-AIP

| **All ages** | **All ED injuries** | **Head injuries** | **TBI** | **% head all ED** | **% TBI all ED** | **% TBI head** |
| --- | --- | --- | --- | --- | --- | --- |
| **2001** | 430,254 | 60,172 | 34,879 | 14.0 | 8.1 | 58.0 |
| **2002** | 404,538 | 54,317 | 30,885 | 13.4 | 7.6 | 56.9 |
| **2003** | 405,961 | 55,202 | 33,505 | 13.6 | 8.3 | 60.7 |
| **2004** | 410,586 | 58,783 | 35,796 | 14.3 | 8.7 | 60.9 |
| **2005** | 373,146 | 53,152 | 33,242 | 14.2 | 8.9 | 62.5 |
| **2006** | 369,892 | 53,649 | 34,626 | 14.5 | 9.4 | 64.5 |
| **2007** | 393,692 | 54,487 | 35,619 | 13.8 | 9.0 | 65.4 |
| **2008** | 395,451 | 60,335 | 39,249 | 15.3 | 9.9 | 65.1 |
| **2009** | 417,986 | 72,604 | 48,809 | 17.4 | 11.7 | 67.2 |
| **2010** | 415,342 | 70,629 | 51,263 | 17.0 | 12.3 | 72.6 |
| **2011** | 419,893 | 70,143 | 53,009 | 16.7 | 12.6 | 75.6 |
| **2012** | 424,819 | 71,493 | 56,618 | 16.8 | 13.3 | 79.2 |
| **2013** | 398,107 | 70,520 | 56,752 | 17.7 | 14.3 | 80.5 |
| **2014** | 391,223 | 67,897 | 52,482 | 17.4 | 13.4 | 77.3 |
| **2015** | 378,099 | 69,283 | 55,068 | 18.3 | 14.6 | 79.5 |
| **2016** | 357,775 | 60,419 | 46,882 | 16.9 | 13.1 | 77.6 |
| **2017** | 340,951 | 56,671 | 43,738 | 16.6 | 12.8 | 77.2 |
| **2018** | 312,511 | 53,676 | 43,164 | 17.2 | 13.8 | 80.4 |
| **2019** | 318,239 | 59,806 | 47,638 | 18.8 | 15.0 | 79.7 |
| **2020** | 318,243 | 66,161 | 53,778 | 20.8 | 16.9 | 81.3 |
| **% changes 2001-10 to 2011-20** |  |  |  |  |  |  |
|  | -8.9% | +8.9% | +34.7% | +20.1% | +48.7% | +24.4% |

**Table 3:** United States cyclist total injury, head only and traumatic brain injury emergency department presentations, 0-17yo, 2001-2020. Source: NEISS-AIP

| **0-17yo** | **All ED injuries** | **Head injuries** | **TBI** | **% head all ED** | **% TBI all ED** | **% TBI head** |
| --- | --- | --- | --- | --- | --- | --- |
| **2001** | 265,614 | 41,989 | 23,441 | 15.8 | 8.8 | 55.8 |
| **2002** | 255,380 | 39,388 | 21,234 | 15.4 | 8.3 | 53.9 |
| **2003** | 257,097 | 37,474 | 22,403 | 14.6 | 8.7 | 59.8 |
| **2004** | 257,637 | 39,948 | 22,538 | 15.5 | 8.7 | 56.4 |
| **2005** | 224,585 | 34,923 | 20,927 | 15.6 | 9.3 | 59.9 |
| **2006** | 209,658 | 32,828 | 18,166 | 15.7 | 8.7 | 55.3 |
| **2007** | 219,249 | 32,782 | 19,690 | 15.0 | 9.0 | 60.1 |
| **2008** | 207,327 | 34,616 | 20,798 | 16.7 | 10.0 | 60.1 |
| **2009** | 223,558 | 40,767 | 25,990 | 18.2 | 11.6 | 63.8 |
| **2010** | 214,253 | 40,611 | 27,684 | 19.0 | 12.9 | 68.2 |
| **2011** | 206,186 | 36,334 | 25,969 | 17.6 | 12.6 | 71.5 |
| **2012** | 202,514 | 34,475 | 25,582 | 17.0 | 12.6 | 74.2 |
| **2013** | 181,215 | 32,547 | 24,945 | 18.0 | 13.8 | 76.6 |
| **2014** | 177,274 | 30,882 | 22,134 | 17.4 | 12.5 | 71.7 |
| **2015** | 156,457 | 27,622 | 20,376 | 17.7 | 13.0 | 73.8 |
| **2016** | 147,296 | 25,011 | 18,516 | 17.0 | 12.6 | 74.0 |
| **2017** | 135,163 | 23,659 | 16,969 | 17.5 | 12.6 | 71.7 |
| **2018** | 117,994 | 21,328 | 16,537 | 18.1 | 14.0 | 77.5 |
| **2019** | 120,357 | 21,795 | 16,740 | 18.1 | 13.9 | 76.8 |
| **2020** | 120,996 | 24,140 | 18,900 | 20.0 | 15.6 | 78.3 |
| **% changes 2001-10 to 2011-20** |  |  |  |  |  |  |
|  | -32.9% | -26.0% | -7.3% | +10.6% | +38.5% | +25.8% |

**Table 4:** United States cyclist total injury, head only and traumatic brain injury emergency department presentations, 18yo+, 2001-2020. Source: NEISS-AIP

| **18yo+** | **All ED injuries** | **Head injuries** | **TBI** | **% head all ED** | **% TBI all ED** | **% TBI head** |
| --- | --- | --- | --- | --- | --- | --- |
| **2001** | 164,640 | 18,183 | * | 11.0 | * | * |
| **2002** | 149,158 | 14,929 | 9,651 | 10.0 | 6.5 | 64.6 |
| **2003** | 148,864 | 17,728 | 11,103 | 11.9 | 7.5 | 62.6 |
| **2004** | 152,949 | 18,835 | 13,242 | 12.3 | 8.7 | 70.3 |
| **2005** | 148,561 | 18,229 | 12,316 | 12.3 | 8.3 | 67.6 |
| **2006** | 160,234 | 20,821 | 16,461 | 13.0 | 10.3 | 79.1 |
| **2007** | 174,443 | 21,705 | 15,929 | 12.4 | 9.1 | 73.4 |
| **2008** | 188,124 | 25,719 | 18,410 | 13.7 | 9.8 | 71.6 |
| **2009** | 194,428 | 31,838 | 22,819 | 16.4 | 11.7 | 71.7 |
| **2010** | 201,089 | 30,018 | 23,579 | 14.9 | 11.7 | 78.5 |
| **2011** | 213,708 | 33,810 | 27,040 | 15.8 | 12.7 | 80.0 |
| **2012** | 222,305 | 37,018 | 31,037 | 16.7 | 14.0 | 83.8 |
| **2013** | 216,892 | 37,973 | 31,807 | 17.5 | 14.7 | 83.8 |
| **2014** | 213,949 | 37,014 | 30,348 | 17.3 | 14.2 | 82.0 |
| **2015** | 221,642 | 41,662 | 34,693 | 18.8 | 15.7 | 83.3 |
| **2016** | 210,479 | 35,408 | 28,366 | 16.8 | 13.5 | 80.1 |
| **2017** | 205,788 | 33,011 | 26,769 | 16.0 | 13.0 | 81.1 |
| **2018** | 194,518 | 32,349 | 26,626 | 16.6 | 13.7 | 82.3 |
| **2019** | 197,883 | 38,010 | 30,898 | 19.2 | 15.6 | 81.3 |
| **2020** | 197,247 | 42,021 | 34,822 | 21.3 | 17.7 | 82.9 |
| **% changes 2001-10 to 2011-20** |  |  |  |  |  |  |
|  | +24.5% | +68.9% | +89.6% | +37.6% | +56.2% | +15.6% |
| * signifies less than 1,200 or Coefficient of Variation exceeds 33% |  |  |  |  |  |  |

**Table 5:** United States cyclist lower and upper body injury emergency department presentations, all-ages, 2001-2020. Source: NEISS-AIP

| **All ages** | **All ED injuries** | **Lower body injuries** | **Upper body injuries** | **% lower body all ED** | **% upper body all ED** |
| --- | --- | --- | --- | --- | --- |
| **2001** | 430,254 | 153,297 | 216,802 | 35.6 | 50.4 |
| **2002** | 404,538 | 146,345 | 203,876 | 36.2 | 50.4 |
| **2003** | 405,961 | 142,042 | 208,717 | 35.0 | 51.4 |
| **2004** | 410,586 | 144,160 | 207,658 | 35.1 | 50.6 |
| **2005** | 373,146 | 133,197 | 186,812 | 35.7 | 50.1 |
| **2006** | 369,892 | 133,159 | 183,085 | 36.0 | 49.5 |
| **2007** | 393,692 | 142,638 | 196,567 | 36.2 | 49.9 |
| **2008** | 395,451 | 145,810 | 189,321 | 36.9 | 47.9 |
| **2009** | 417,986 | 144,898 | 200,484 | 34.7 | 48.0 |
| **2010** | 415,342 | 147,729 | 196,984 | 35.6 | 47.4 |
| **2011** | 419,893 | 149,494 | 200,256 | 35.6 | 47.7 |
| **2012** | 424,819 | 153,565 | 199,761 | 36.1 | 47.0 |
| **2013** | 398,107 | 142,032 | 185,555 | 35.7 | 46.6 |
| **2014** | 391,223 | 141,440 | 181,887 | 36.2 | 46.5 |
| **2015** | 378,099 | 134,082 | 174,733 | 35.5 | 46.2 |
| **2016** | 357,775 | 130,954 | 166,418 | 36.6 | 46.5 |
| **2017** | 340,951 | 122,813 | 161,468 | 36.0 | 47.4 |
| **2018** | 312,511 | 111,895 | 146,940 | 35.8 | 47.0 |
| **2019** | 318,239 | 128,788 | 168,762 | 40.5 | 53.0 |
| **2020** | 318,243 | 121,392 | 174,955 | 38.1 | 55.0 |
| **% changes 2001-10 to 2011-20** |  |  |  |  |  |
|  | -8.9% | -6.8% | -11.5% | +2.5% | -2.6% |
| Lower body = pubic region, lower trunk, knee, lower leg, ankle, upper leg, foot, toe |  |  |  |  |  |
| Upper body = shoulder, elbow, lower arm, wrist, upper arm, hand, finger, upper trunk |  |  |  |  |  |

**Table 6:**
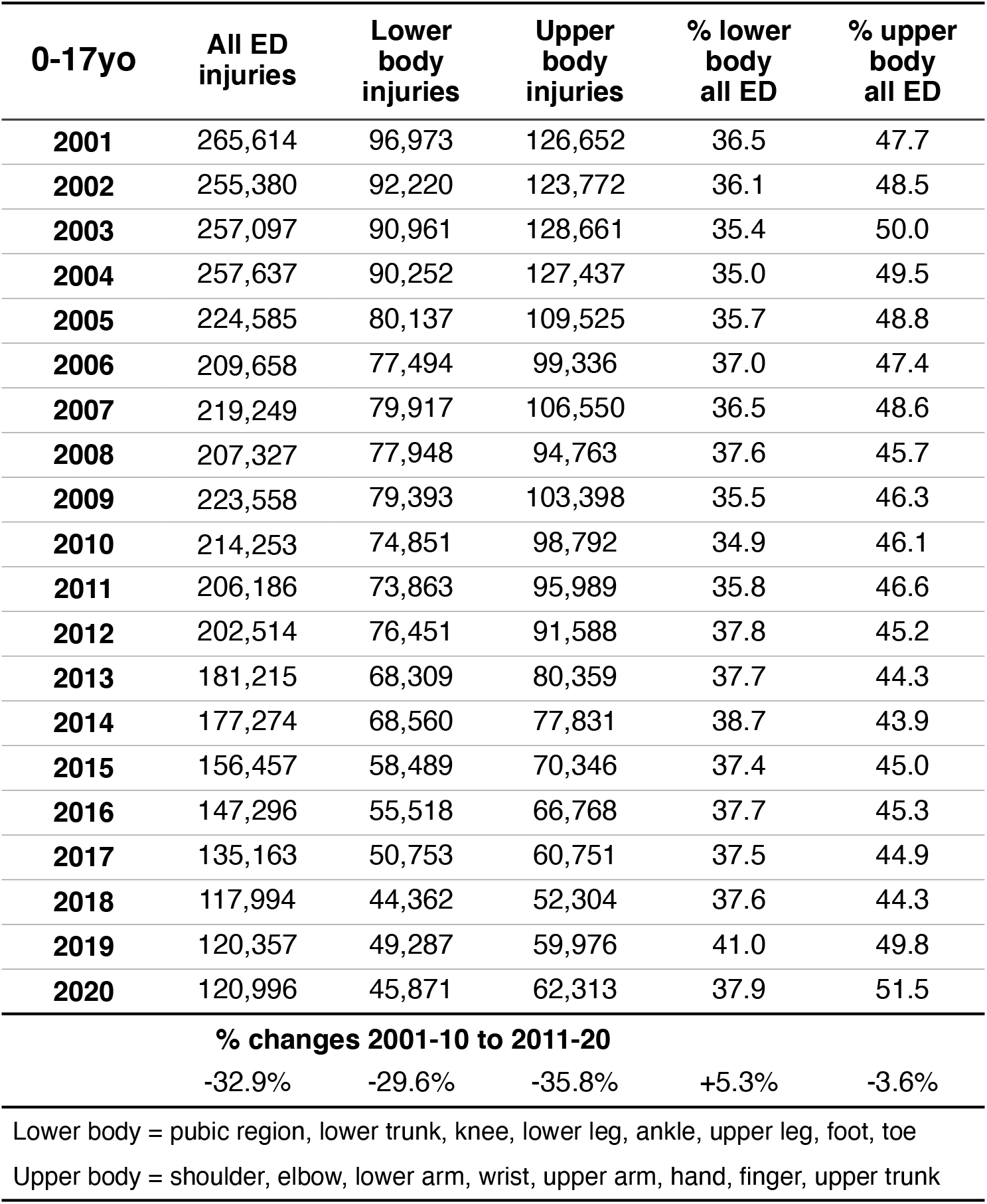
United States cyclist lower and upper body injury emergency department presentations, 0-17yo, 2001-2020. Source: NEISS-AIP

| **0-17yo** | **All ED injuries** | **Lower body injuries** | **Upper body injuries** | **% lower body all ED** | **% upper body all ED** |
| --- | --- | --- | --- | --- | --- |
| **2001** | 265,614 | 96,973 | 126,652 | 36.5 | 47.7 |
| **2002** | 255,380 | 92,220 | 123,772 | 36.1 | 48.5 |
| **2003** | 257,097 | 90,961 | 128,661 | 35.4 | 50.0 |
| **2004** | 257,637 | 90,252 | 127,437 | 35.0 | 49.5 |
| **2005** | 224,585 | 80,137 | 109,525 | 35.7 | 48.8 |
| **2006** | 209,658 | 77,494 | 99,336 | 37.0 | 47.4 |
| **2007** | 219,249 | 79,917 | 106,550 | 36.5 | 48.6 |
| **2008** | 207,327 | 77,948 | 94,763 | 37.6 | 45.7 |
| **2009** | 223,558 | 79,393 | 103,398 | 35.5 | 46.3 |
| **2010** | 214,253 | 74,851 | 98,792 | 34.9 | 46.1 |
| **2011** | 206,186 | 73,863 | 95,989 | 35.8 | 46.6 |
| **2012** | 202,514 | 76,451 | 91,588 | 37.8 | 45.2 |
| **2013** | 181,215 | 68,309 | 80,359 | 37.7 | 44.3 |
| **2014** | 177,274 | 68,560 | 77,831 | 38.7 | 43.9 |
| **2015** | 156,457 | 58,489 | 70,346 | 37.4 | 45.0 |
| **2016** | 147,296 | 55,518 | 66,768 | 37.7 | 45.3 |
| **2017** | 135,163 | 50,753 | 60,751 | 37.5 | 44.9 |
| **2018** | 117,994 | 44,362 | 52,304 | 37.6 | 44.3 |
| **2019** | 120,357 | 49,287 | 59,976 | 41.0 | 49.8 |
| **2020** | 120,996 | 45,871 | 62,313 | 37.9 | 51.5 |
| **% changes 2001-10 to 2011-20** |  |  |  |  |  |
|  | -32.9% | -29.6% | -35.8% | +5.3% | -3.6% |
| Lower body = pubic region, lower trunk, knee, lower leg, ankle, upper leg, foot, toe |  |  |  |  |  |
| Upper body = shoulder, elbow, lower arm, wrist, upper arm, hand, finger, upper trunk |  |  |  |  |  |

**Table 7:** United States cyclist lower and upper body injury emergency department presentations, 18yo+, 2001-2020. Source: NEISS-AIP

| **18yo+** | **All ED injuries** | **Lower body injuries** | **Upper body injuries** | **% lower body all ED** | **% upper body all ED** |
| --- | --- | --- | --- | --- | --- |
| **2001** | 164,640 | 56,324 | 90,133 | 34.2 | 54.7 |
| **2002** | 149,158 | 54,125 | 80,104 | 36.3 | 53.7 |
| **2003** | 148,864 | 51,080 | 80,055 | 34.3 | 53.8 |
| **2004** | 152,949 | 53,909 | 80,206 | 35.2 | 52.4 |
| **2005** | 148,561 | 53,060 | 77,272 | 35.7 | 52.0 |
| **2006** | 160,234 | 55,665 | 83,748 | 34.7 | 52.3 |
| **2007** | 174,443 | 62,721 | 90,017 | 36.0 | 51.6 |
| **2008** | 188,124 | 67,862 | 94,543 | 36.1 | 50.3 |
| **2009** | 194,428 | 65,505 | 97,085 | 33.7 | 49.9 |
| **2010** | 201,089 | 72,878 | 98,192 | 36.2 | 48.8 |
| **2011** | 213,708 | 75,631 | 104,267 | 35.4 | 48.8 |
| **2012** | 222,305 | 77,113 | 108,173 | 34.7 | 48.7 |
| **2013** | 216,892 | 73,723 | 105,196 | 34.0 | 48.5 |
| **2014** | 213,949 | 72,879 | 104,056 | 34.1 | 48.6 |
| **2015** | 221,642 | 75,593 | 104,387 | 34.1 | 47.1 |
| **2016** | 210,479 | 75,436 | 99,635 | 35.8 | 47.3 |
| **2017** | 205,788 | 72,060 | 100,717 | 35.0 | 48.9 |
| **2018** | 194,518 | 67,533 | 94,635 | 34.7 | 48.7 |
| **2019** | 197,883 | 79,502 | 108,786 | 40.2 | 55.0 |
| **2020** | 197,247 | 75,521 | 112,642 | 38.3 | 57.1 |
| **% changes 2001-10 to 2011-20** |  |  |  |  |  |
|  | +24.5% | +25.6% | +19.6% | +1.1% | -4.0% |
| Lower body = pubic region, lower trunk, knee, lower leg, ankle, upper leg, foot, toe |  |  |  |  |  |
| Upper body = shoulder, elbow, lower arm, wrist, upper arm, hand, finger, upper trunk |  |  |  |  |  |

**Table 8:**
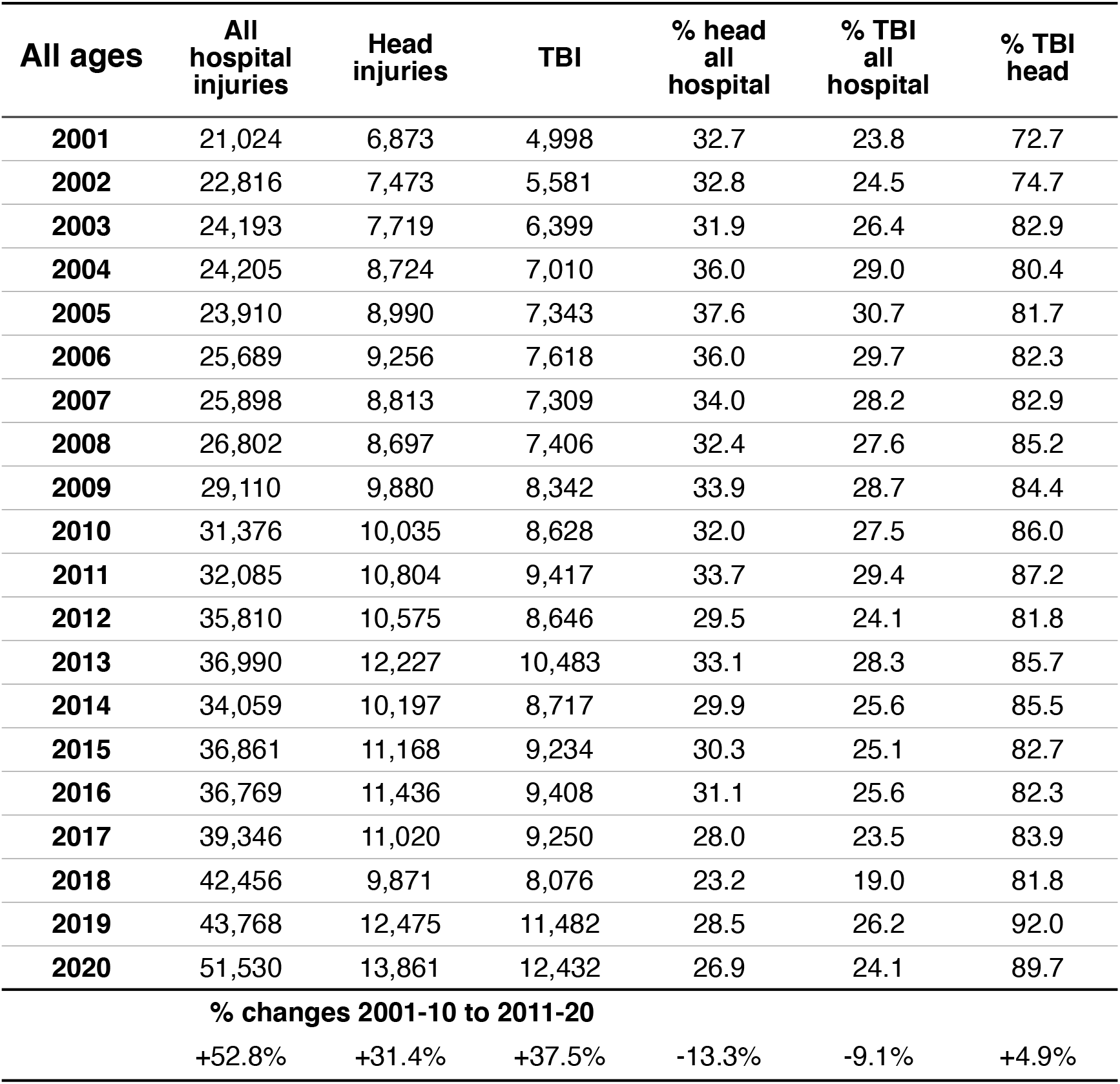
United States cyclist total injury, head only and traumatic brain injury hospital admissions, all-ages, 2001-2020. Source: NEISS-AIP

| **All ages** | **All hospital injuries** | **Head injuries** | **TBI** | **% head all hospital** | **% TBI all hospital** | **% TBI head** |
| --- | --- | --- | --- | --- | --- | --- |
| **2001** | 21,024 | 6,873 | 4,998 | 32.7 | 23.8 | 72.7 |
| **2002** | 22,816 | 7,473 | 5,581 | 32.8 | 24.5 | 74.7 |
| **2003** | 24,193 | 7,719 | 6,399 | 31.9 | 26.4 | 82.9 |
| **2004** | 24,205 | 8,724 | 7,010 | 36.0 | 29.0 | 80.4 |
| **2005** | 23,910 | 8,990 | 7,343 | 37.6 | 30.7 | 81.7 |
| **2006** | 25,689 | 9,256 | 7,618 | 36.0 | 29.7 | 82.3 |
| **2007** | 25,898 | 8,813 | 7,309 | 34.0 | 28.2 | 82.9 |
| **2008** | 26,802 | 8,697 | 7,406 | 32.4 | 27.6 | 85.2 |
| **2009** | 29,110 | 9,880 | 8,342 | 33.9 | 28.7 | 84.4 |
| **2010** | 31,376 | 10,035 | 8,628 | 32.0 | 27.5 | 86.0 |
| **2011** | 32,085 | 10,804 | 9,417 | 33.7 | 29.4 | 87.2 |
| **2012** | 35,810 | 10,575 | 8,646 | 29.5 | 24.1 | 81.8 |
| **2013** | 36,990 | 12,227 | 10,483 | 33.1 | 28.3 | 85.7 |
| **2014** | 34,059 | 10,197 | 8,717 | 29.9 | 25.6 | 85.5 |
| **2015** | 36,861 | 11,168 | 9,234 | 30.3 | 25.1 | 82.7 |
| **2016** | 36,769 | 11,436 | 9,408 | 31.1 | 25.6 | 82.3 |
| **2017** | 39,346 | 11,020 | 9,250 | 28.0 | 23.5 | 83.9 |
| **2018** | 42,456 | 9,871 | 8,076 | 23.2 | 19.0 | 81.8 |
| **2019** | 43,768 | 12,475 | 11,482 | 28.5 | 26.2 | 92.0 |
| **2020** | 51,530 | 13,861 | 12,432 | 26.9 | 24.1 | 89.7 |
| **% changes 2001-10 to 2011-20** |  |  |  |  |  |  |
|  | +52.8% | +31.4% | +37.5% | -13.3% | -9.1% | +4.9% |

**Table 9:** United States cyclist total injury, head only and traumatic brain injury hospital admissions, 0-17yo, 2001-2020. Source: NEISS-AIP

| **0-17yo** | **All hospital injuries** | **Head injuries** | **TBI** | **% head all hospital** | **% TBI all hospital** | **% TBI head** |
| --- | --- | --- | --- | --- | --- | --- |
| **2001** | 10,969 | 3,776 | 2,734 | 34.4 | 24.9 | 72.4 |
| **2002** | 11,815 | 3,764 | 2,681 | 31.9 | 22.7 | 71.2 |
| **2003** | 13,121 | 4,193 | 3,612 | 32.0 | 27.5 | 86.1 |
| **2004** | 12,226 | 5,012 | 4,024 | 41.0 | 32.9 | 80.3 |
| **2005** | 11,033 | 4,027 | 3,112 | 36.5 | 28.2 | 77.3 |
| **2006** | 11,234 | 4,352 | 3,355 | 38.7 | 29.9 | 77.1 |
| **2007** | 10,121 | 3,411 | 2,733 | 33.7 | 27.0 | 80.1 |
| **2008** | 9,454 | 3,653 | 2,750 | 38.6 | 29.1 | 75.3 |
| **2009** | 9,753 | 3,589 | 2,947 | 36.8 | 30.2 | 82.1 |
| **2010** | 9,307 | 2,945 | 2,495 | 31.6 | 26.8 | 84.7 |
| **2011** | 10,509 | 3,606 | 2,949 | 34.3 | 28.1 | 81.8 |
| **2012** | 10,094 | 2,656 | 1,740 | 26.3 | 17.2 | 65.5 |
| **2013** | 8,063 | 2,433 | 2,099 | 30.2 | 26.0 | 86.3 |
| **2014** | 8,278 | 2,387 | 1,973 | 28.8 | 23.8 | 82.7 |
| **2015** | 7,707 | 3,371 | 2,629 | 43.7 | 34.1 | 78.0 |
| **2016** | 7,448 | 2,633 | 1,879 | 35.4 | 25.2 | 71.4 |
| **2017** | 6,448 | 1,402 | * | 21.7 | * | * |
| **2018** | 6,244 | 1,733 | 1,578 | 27.8 | 25.3 | 91.1 |
| **2019** | 6,563 | 1,808 | 1,692 | 27.5 | 25.8 | 93.6 |
| **2020** | 9,635 | 2,572 | 2,147 | 26.7 | 22.3 | 83.5 |
| **% changes 2001-10 to 2011-20** |  |  |  |  |  |  |
|  | -25.7% | -36.5% | -31.8% | -14.9% | -9.3% | +3.6% |
| * signifies less than 1,200 or Coefficient of Variation exceeds 33% |  |  |  |  |  |  |

**Table 10:** United States cyclist total injury, head only and traumatic brain injury hospital admissions, 18yo+, 2001-2020. Source: NEISS-AIP

| **18yo+** | **All hospital injuries** | **Head injuries** | **TBI** | **% head all hospital** | **% TBI all hospital** | **% TBI head** |
| --- | --- | --- | --- | --- | --- | --- |
| **2001** | 10,056 | 3,097 | 2,264 | 30.8 | 22.5 | 73.1 |
| **2002** | 11,001 | * | * | * | * | * |
| **2003** | 11,072 | 3,526 | 2,788 | 31.8 | 25.2 | 79.1 |
| **2004** | 11,979 | 3,712 | 2,986 | 31.0 | 24.9 | 80.4 |
| **2005** | 12,877 | 4,948 | * | 38.4 | * | * |
| **2006** | 14,455 | * | * | * | * | * |
| **2007** | 15,777 | 5,402 | 4,577 | 34.2 | 29.0 | 84.7 |
| **2008** | 17,348 | 5,044 | 4,655 | 29.1 | 26.8 | 92.3 |
| **2009** | 19,357 | 6,292 | 5,396 | 32.5 | 27.9 | 85.8 |
| **2010** | 22,068 | 7,089 | 6,133 | 32.1 | 27.8 | 86.5 |
| **2011** | 21,575 | 7,182 | 6,452 | 33.3 | 29.9 | 89.8 |
| **2012** | 25,716 | 7,919 | 6,906 | 30.8 | 26.9 | 87.2 |
| **2013** | 28,927 | 9,717 | 8,306 | 33.6 | 28.7 | 85.5 |
| **2014** | 25,781 | 7,778 | 6,713 | 30.2 | 26.0 | 86.3 |
| **2015** | 29,154 | 7,782 | 6,590 | 26.7 | 22.6 | 84.7 |
| **2016** | 29,321 | 8,803 | 7,529 | 30.0 | 25.7 | 85.5 |
| **2017** | 32,898 | 9,618 | 8,150 | 29.2 | 24.8 | 84.7 |
| **2018** | 36,212 | 8,138 | 6,498 | 22.5 | 17.9 | 79.8 |
| **2019** | 37,205 | 10,551 | 9,675 | 28.4 | 26.0 | 91.7 |
| **2020** | 41,894 | 11,280 | 10,275 | 26.9 | 24.5 | 91.1 |
| **% changes 2001-10 to 2011-20** |  |  |  |  |  |  |
|  | +111.4% | +81.6% | +87.4% | -10.2% | -3.8% | +4.2% |
| * signifies less than 1,200 or Coefficient of Variation exceeds 33% |  |  |  |  |  |  |

**Table 11:** United States cyclist lower and upper body injury hospital admissions, all-ages, 2001-2020. Source: NEISS-AIP

| **All ages** | **All hospital injuries** | **Lower body injuries** | **Upper body injuries** | **% lower body all hospital** | **% upper body all hospital** |
| --- | --- | --- | --- | --- | --- |
| **2001** | 21,024 | 7,406 | 6,745 | 35.2 | 32.1 |
| **2002** | 22,816 | 8,087 | 7,256 | 35.4 | 31.8 |
| **2003** | 24,193 | 8,123 | 8,350 | 33.6 | 34.5 |
| **2004** | 24,205 | 8,845 | 6,635 | 36.5 | 27.4 |
| **2005** | 23,910 | 7,427 | 7,508 | 31.1 | 31.4 |
| **2006** | 25,689 | 9,010 | 7,489 | 35.1 | 29.2 |
| **2007** | 25,898 | 8,577 | 8,507 | 33.1 | 32.8 |
| **2008** | 26,802 | 8,768 | 9,337 | 32.7 | 34.8 |
| **2009** | 29,110 | 9,798 | 9,432 | 33.7 | 32.4 |
| **2010** | 31,376 | 9,911 | 11,430 | 31.6 | 36.4 |
| **2011** | 32,085 | 10,558 | 10,739 | 32.9 | 33.5 |
| **2012** | 35,810 | 11,601 | 13,650 | 32.4 | 38.1 |
| **2013** | 36,990 | 11,623 | 13,218 | 31.4 | 35.7 |
| **2014** | 34,059 | 10,297 | 13,597 | 30.2 | 39.9 |
| **2015** | 36,861 | 11,723 | 13,985 | 31.8 | 37.9 |
| **2016** | 36,769 | 12,498 | 12,850 | 34.0 | 34.9 |
| **2017** | 39,346 | 11,689 | 16,637 | 29.7 | 42.3 |
| **2018** | 42,456 | 15,045 | 17,540 | 35.4 | 41.3 |
| **2019** | 43,768 | 17,494 | 21,909 | 40.0 | 50.1 |
| **2020** | 51,530 | 21,400 | 26,173 | 41.5 | 50.8 |
| **% changes 2001-10 to 2011-20** |  |  |  |  |  |
|  | +52.8% | +55.8% | +93.9% | +0.3% | +25.4% |
| Lower body = pubic region, lower trunk, knee, lower leg, ankle, upper leg, foot, toe |  |  |  |  |  |
| Upper body = shoulder, elbow, lower arm, wrist, upper arm, hand, finger, upper trunk |  |  |  |  |  |

**Table 12:** United States cyclist lower and upper body injury hospital admissions, 0-17yo, 2001-2020. Source: NEISS-AIP

| **0-17yo** | **All hospital injuries** | **Lower body injuries** | **Upper body injuries** | **% lower body all hospital** | **% upper body all hospital** |
| --- | --- | --- | --- | --- | --- |
| **2001** | 10,969 | 3,253 | 3,940 | 29.7 | 35.9 |
| **2002** | 11,815 | 3,642 | 4,410 | 30.8 | 37.3 |
| **2003** | 13,121 | 4,010 | 4,917 | 30.6 | 37.5 |
| **2004** | 12,226 | 3,334 | 3,880 | 27.3 | 31.7 |
| **2005** | 11,033 | 2,996 | 4,010 | 27.2 | 36.3 |
| **2006** | 11,234 | 3,827 | 3,055 | 34.1 | 27.2 |
| **2007** | 10,121 | 2,568 | 4,141 | 25.4 | 40.9 |
| **2008** | 9,454 | 2,437 | 3,364 | 25.8 | 35.6 |
| **2009** | 9,753 | 2,835 | 3,329 | 29.1 | 34.1 |
| **2010** | 9,307 | 2,627 | 3,735 | 28.2 | 40.1 |
| **2011** | 10,509 | 2,950 | 3,953 | 28.1 | 37.6 |
| **2012** | 10,094 | 3,655 | 3,783 | 36.2 | 37.5 |
| **2013** | 8,063 | 2,554 | 3,076 | 31.7 | 38.1 |
| **2014** | 8,278 | 2,579 | 3,311 | 31.2 | 40.0 |
| **2015** | 7,707 | 1,972 | 2,364 | 25.6 | 30.7 |
| **2016** | 7,448 | 1,835 | 2,980 | 24.6 | 40.0 |
| **2017** | 6,448 | 2,057 | 2,989 | 31.9 | 46.4 |
| **2018** | 6,244 | 2,053 | 2,458 | 32.9 | 39.4 |
| **2019** | 6,563 | 2,521 | 3,450 | 38.4 | 52.6 |
| **2020** | 9,635 | 3,542 | 4,763 | 36.8 | 49.4 |
| **% changes 2001-10 to 2011-20** |  |  |  |  |  |
|  | -25.7% | -18.4% | -14.6% | +10.1% | +15.4% |
| Lower body = pubic region, lower trunk, knee, lower leg, ankle, upper leg, foot, toe |  |  |  |  |  |
| Upper body = shoulder, elbow, lower arm, wrist, upper arm, hand, finger, upper trunk |  |  |  |  |  |

**Table 13:**
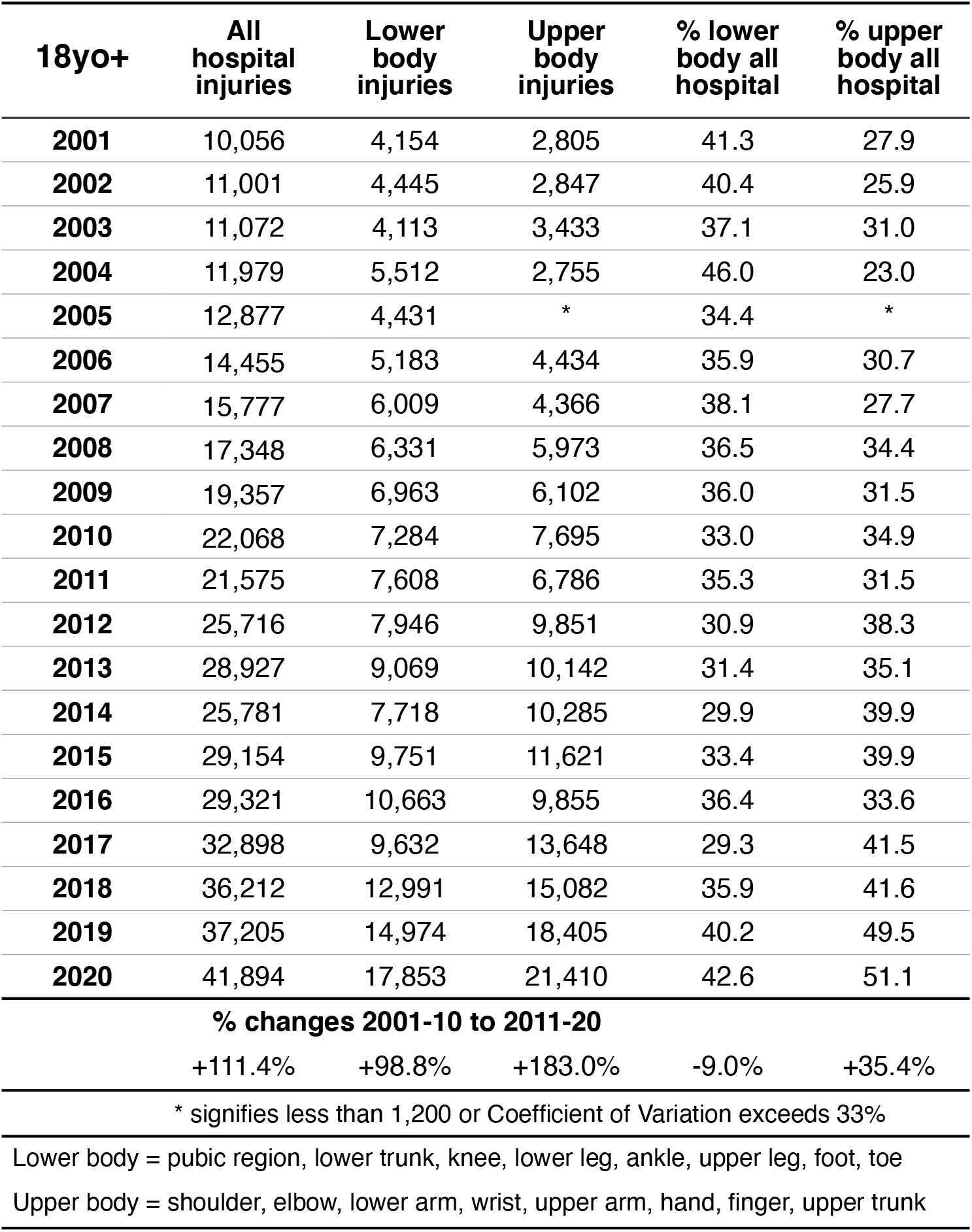
United States cyclist lower and upper body injury hospital admissions, 18yo+, 2001-2020. Source: NEISS-AIP

| **18yo+** | **All hospital injuries** | **Lower body injuries** | **Upper body injuries** | **% lower body all hospital** | **% upper body all hospital** |
| --- | --- | --- | --- | --- | --- |
| **2001** | 10,056 | 4,154 | 2,805 | 41.3 | 27.9 |
| **2002** | 11,001 | 4,445 | 2,847 | 40.4 | 25.9 |
| **2003** | 11,072 | 4,113 | 3,433 | 37.1 | 31.0 |
| **2004** | 11,979 | 5,512 | 2,755 | 46.0 | 23.0 |
| **2005** | 12,877 | 4,431 | * | 34.4 | * |
| **2006** | 14,455 | 5,183 | 4,434 | 35.9 | 30.7 |
| **2007** | 15,777 | 6,009 | 4,366 | 38.1 | 27.7 |
| **2008** | 17,348 | 6,331 | 5,973 | 36.5 | 34.4 |
| **2009** | 19,357 | 6,963 | 6,102 | 36.0 | 31.5 |
| **2010** | 22,068 | 7,284 | 7,695 | 33.0 | 34.9 |
| **2011** | 21,575 | 7,608 | 6,786 | 35.3 | 31.5 |
| **2012** | 25,716 | 7,946 | 9,851 | 30.9 | 38.3 |
| **2013** | 28,927 | 9,069 | 10,142 | 31.4 | 35.1 |
| **2014** | 25,781 | 7,718 | 10,285 | 29.9 | 39.9 |
| **2015** | 29,154 | 9,751 | 11,621 | 33.4 | 39.9 |
| **2016** | 29,321 | 10,663 | 9,855 | 36.4 | 33.6 |
| **2017** | 32,898 | 9,632 | 13,648 | 29.3 | 41.5 |
| **2018** | 36,212 | 12,991 | 15,082 | 35.9 | 41.6 |
| **2019** | 37,205 | 14,974 | 18,405 | 40.2 | 49.5 |
| **2020** | 41,894 | 17,853 | 21,410 | 42.6 | 51.1 |
| **% changes 2001-10 to 2011-20** |  |  |  |  |  |
|  | +111.4% | +98.8% | +183.0% | -9.0% | +35.4% |
| * signifies less than 1,200 or Coefficient of Variation exceeds 33% |  |  |  |  |  |
| Lower body = pubic region, lower trunk, knee, lower leg, ankle, upper leg, foot, toe |  |  |  |  |  |
| Upper body = shoulder, elbow, lower arm, wrist, upper arm, hand, finger, upper trunk |  |  |  |  |  |

**Table 14:** United States average hospitalized cyclist, vehicle occupant and pedestrian injuries in 2001-2009 and 2011-2019. Source: WISQARS

| **5-15yo** | **Pedal cyclists** |  | **Vehicle occupants** |  | **Pedestrians** |  |
| --- | --- | --- | --- | --- | --- | --- |
|  | Average number | Crude rate per 100,000 | Average number | Crude rate per 100,000 | Average number | Crude rate per 100,000 |
| **2001-2009** | 6,519 | 14.45 | 10,097 | 22.34 | 3,955 | 8.76 |
| **2011-2019** | 3,769 | 8.32 | 4,580 | 10.11 | 2,956 | 6.52 |
| **% change** | 57.8% | 57.6% | 45.4% | 45.2% | 74.8% | 74.5% |

| **16yo+** | **Pedal cyclists** |  | **Vehicle occupants** |  | **Pedestrians** |  |
| --- | --- | --- | --- | --- | --- | --- |
|  | Average number | Crude rate per 100,000 | Average number | Crude rate per 100,000 | Average number | Crude rate per 100,000 |
| **2001-2009** | 13,690 | 5.91 | 165,638 | 72.13 | 21,862 | 9.36 |
| **2011-2019** | 27,276 | 10.65 | 170,753 | 66.78 | 37,168 | 14.53 |
| **% change** | 199.2% | 180.2% | 103.1% | 92.6% | 170.0% | 155.2% |

Source : Centers for Disease Control and Prevention. Web-based Injury Statistics Query and Reporting System (WISQARS) [Online]. (2003). National Center for Injury Prevention and Control, Centers for Disease Control and Prevention (producer). Available from: URL: www.cdc.gov/injury/wisqars. 2021/10/15

**Table 15:** United States 6-17yo average cycling participation and hospitalized total and head injuries. Source: OIA and NEISS

| **6-17yo** | **OIA cycling participation** | **NEISS total injuries** | **Total injuries per 100,000** | **Head injuries** | **Head injuries per 100,000** |
| --- | --- | --- | --- | --- | --- |
| **2006** | 17,463,000 | 10,340 | 59.21 | 3,698 | 21.18 |
| **2007** | 15,550,000 | 9,981 | 64.19 | 3,170 | 20.39 |
| **2008** | 14,716,000 | 9,060 | 61.57 | 3,297 | 22.40 |
| **2009** | 14,652,000 | 9,610 | 65.59 | 3,264 | 22.28 |
| **2010** | 13,657,000 | 9,069 | 66.40 | 2,809 | 20.57 |
| **2011** | 13,283,000 | 10,559 | 79.50 | 3,520 | 26.50 |
| **2012** | 13,421,000 | 10,263 | 76.47 | 2,395 | 17.85 |
| **2013** | 13,498,000 | 8,205 | 60.79 | 2,264 | 16.77 |
| **2104** | 12,953,000 | 8,693 | 67.11 | 2,220 | 17.14 |
| **2015** | 12,461,000 | 7,626 | 61.20 | 2,819 | 22.62 |
| **2016** | 12,889,000 | 6,977 | 54.13 | 2,281 | 17.70 |
| **2017** | 12,535,000 | 6,338 | 50.56 | * | * |
| **2018** | 12,703,000 | 6,295 | 49.56 | 1,549 | 12.19 |
| **2019** | 12,743,000 | 6,390 | 50.15 | 1,738 | 13.64 |
| **2006-2012** | 14,677,429 | 9,840 | 67.56 | 3,165 | 21.59 |
| **2013-2019** | 12,826,000 | 7,218 | 56.21 | 2,145 | 16.68 |
| **% change** | -12.6% | -26.6% | -16.8% | -32.2% | -22.7% |

**Table 16:**
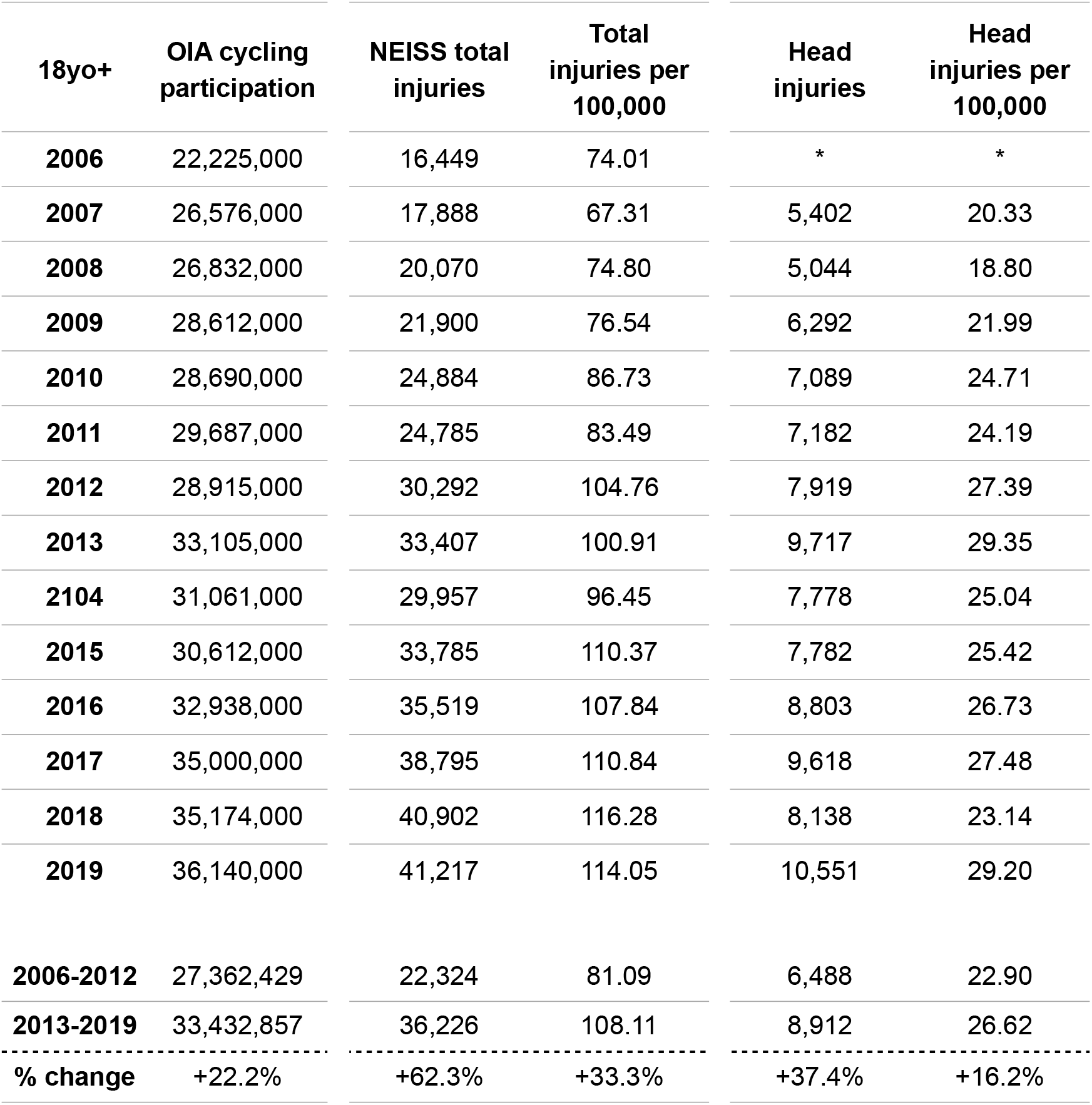
United States 18yo+ average cycling participation and hospitalized total and head injuries. Source: OIA and NEISS

**Table 17:** United States 5-15yo and 16yo+ average cycling participation and hospitalized total and head injuries. Source: NHTS and NEISS *Note*: Head injuries in this table include head, face, eyeballs, mouth, neck and ears

| **5-15yo** | **NHTS**<br>**cycling**<br>**participation** | **NEISS total**<br>**injuries** | **Total**<br>**injuries per**<br>**100,000** | **Head**<br>**injuries** | **Head**<br>**injuries per**<br>**100,000** |
| --- | --- | --- | --- | --- | --- |
| **2001** | 18,560,000 | 10,725 | 57.79 | 3,937 | 21.21 |
| **2009** | 15,920,000 | 9,150 | 57.47 | 3,925 | 24.66 |
| **2017** | 7,410,000 | 6,321 | 85.30 | 1,707 | 23.04 |
| **16yo+** | **NHTS**<br>**cycling**<br>**participation** | **NEISS total**<br>**injuries** | **Total**<br>**injuries per**<br>**100,000** | **Head**<br>**injuries** | **Head**<br>**injuries per**<br>**100,000** |
| **2001** | 14,580,000 | 12,509 | 85.80 | 4,452 | 30.54 |
| **2009** | 24,890,000 | 22,797 | 91.60 | 8,806 | 35.38 |
| **2017** | 28,240,000 | 39,471 | 139.77 | 14,479 | 51.27 |

